# Postacute Sequelae SARS-CoV-2 Infection by Vaccination Status: A Six-Month Latent Class Analysis

**DOI:** 10.1101/2023.10.20.23297332

**Authors:** Xiaowu Sun, Jonathan P. DeShazo, Laura Anatale-Tardiff, Manuela Di Fusco, Kristen E. Allen, Thomas M. Porter, Henriette Coetzer, Santiago M.C Lopez, Laura Puzniak, Joseph C. Cappelleri

## Abstract

Symptoms post-SARS-CoV-2 infection may persist for months and cause significant impairment and impact to quality of life. Acute symptoms of SARS-CoV-2 infection are well studied, yet data on clusters of symptoms over time, or postacute sequelae of SARS-CoV-2 infection (PASC), are limited. We aim to characterize PASC phenotypes by identifying symptom clusters over a six-month period following infection in individuals vaccinated (boosted and not) and those unvaccinated.

Subjects with ≥1 self-reported symptom and positive RT-PCR for SARS-CoV-2 at CVS Health US test sites were recruited between January and April 2022. Patient-reported outcomes symptoms, heath-related quality of life (QoL), work productivity and activity impairment (WPAI) were captured at 1 month, 3 months, and 6 months post-acute infection.

Logistic regression and latent class analysis (LCA) were performed on 20 symptoms using baseline socio-demographic, clinical characteristics, and vaccination status as well as EQ-5and WPAI results as covariables. Subjects with more symptoms were associated with lower health-related quality of life, and worse WPAI scores.

LCA identified three phenotypes that are primarily differentiated by number of symptoms. These three phenotypes remained consistent across time periods. Vaccinated individuals were more likely to be in the low symptom burden latent classes at all time points compared to unvaccinated individuals.

## Introduction & Background

Lingering symptoms of SARS-CoV-2 infection can affect a person’s quality of life and ability to return to work (Di Fusco et al. 2022). New, recurring, and ongoing symptoms may present later than 4 weeks post infection, persist for months, and be similar, or different from, the acute stage symptoms (Centers for Disease Control and Prevention 2022). A systematic literature review found that even after *mild* SARS-COV-2 infection, symptoms persisted more than three weeks in 10% to 35% of patients, and significantly impacted work and daily living (van Kessel et al. 2022). While there is heterogeneity in the identification and definitions surrounding the symptoms and conditions after the acute phase of SAR-CoV-2 infection, in this paper the term PASC is used.

Although data are still limited regarding the underlying pathophysiology of PASC (Nalbandian et al. 2021), understanding the patient symptomatology and care needs after the acute stages of SARS-CoV-2 infection can lead to potential interventions addressing PASC. Furthermore, there are a paucity of data in identifying clusters of symptoms associated with PASC (Natarajan et al. 2023, Priyal et al. 2023, Thaweethai et al. 2023, Yugar-Toledo et al. 2023).

LCA has been an instrumental tool to identify disease phenotypes and targeted therapies. LCA identified acute respiratory distress syndrome (ARDS) phenotypes that respond to distinct treatments (Dahmer et al. 2022, Famous et al. 2017, Calfee et al. 2014). Researchers have also used LCA to identify SARS-CoV-2 phenotypes in patients admitted to the ICU (Sinha et al. 2021, Sigwadhi et al. 2022), describe phenotypes specific to SARS-CoV-2 headaches (Karadaş et al. 2021), predict hospitalization in confirmed SARS-CoV-2 cases (Wang et al. 2021), describe unconfirmed SARS-CoV-2 sequalae in Brazil (Moreira 2021) and Iran, (Hosseinzadeh et al. 2021), and predict SARS-CoV-2 test results based on patterns of symptoms (French et al. 2021). The purpose of this research is to characterize PASC phenotypes by identifying symptom clusters over a six-month period following confirmed infection.

## Methods

We followed best practices for using LCA to identify patient subclasses (Weller, Bowen, and Faubert 2020, Atreya, Sanchez-Pinto, and Kamaleswaran 2022). LCA model selection was based on Bayesian Information Criteria (BIC) and class size prior to applying covariates and interpreting the phenotypes. Finally, we evaluated the model phenotypes through comparison of key covariables.

### Participants

We conducted a secondary data analysis using self-reported outcomes from a previous study (Di Fusco et al. 2022). The data are from symptomatic adults ≥ 18 years old that tested positive for SARS-CoV-2 at one of ∼5,000 CVS Health test sites across the US between 01/31/2022 and 04/30/2022, with six months of follow up. Subjects reported symptoms and other outcomes at three different time points: 4 weeks, 3 months, and 6 months following a SARS-CoV-2 laboratory-confirmed positive test result.

### Measures

The primary data for our LCA were categorical symptoms at each of the three time points. However, we also included vaccination status, activity impairment, and overall quality of life to interpret and validate the model.

#### Post-COVID symptoms

The list of symptoms reflected the CDC’s definition of PASC at that time (Centers for Disease Control and Prevention 2022). Presence of symptoms was reported using a checklist-style survey previously described (Di Fusco et al. 2023).

#### Vaccination status

The study population was classified in three mutually exclusive vaccine cohorts: (1) the “primed” cohort of subjects who received primary series vaccine of BNT162b2 (original monovalent vaccine), (2) the “boosted” cohort of subjects with at least one booster dose after primary series of BNT162b2 (original monovalent vaccine), and (3) the “unvaccinated” cohort of subject with no evidence of vaccination.

#### Activity Impairment

From Work Productivity and Impairment (WPAI) Questionnaire (Reilly, Zbrozek, and Dukes 1993), we selected a single impairment item for use in this analysis: “Consider only how much health problems affected your ability to do your regular daily activities”, elicited using a 0-10 visual analogue scale. The activity impairment score was derived from the item, with range of 0-100% and higher scores indicating more activity impairment (Reilly, Zbrozek, and Dukes 1993).

#### Health-Related Quality of Life

Health-Related Quality of Life (HRQoL) was captured using the EQ-5D-5L questionnaire (EuroQol Research Foundation 2019) and a single utility index was derived using Pickard et. al methods (Pickard et al. 2019).

### Analysis

Analysis was performed using R 4.2.1 and the poLCA_1.6.0.1 package. To identify latent classes of PASC self-reported symptoms, we first estimated models ranging from 1 class to 10 classes. We examined model fit based on our understanding of PASC related symptoms and the following statistical criteria: 1) with lower Bayesian Information Criteria (BIC) indicating better model fit (Nylund, Asparouhov, and Muthén 2007), and 2) Minimizing undesirable small sample sizes (Masyn 2013). Based on these criteria, we identified the best class model as having three classes.

After identifying the best number of classes, we added vaccination status as a covariate and created the final class model. Finally, each subject was then assigned to a specific class for each reporting month based on their posterior class membership (Asparouhov T 2014, Vermunt 2010)

Table 1 presents the relevant characteristics of the study population as well as responses to predictor variables over time.

**Table 1:** Participant characteristics and responses to indicator variables.

|  | **WEEK 4** | **MONTH 3** | **MONTH 6** |
| --- | --- | --- | --- |
| Total, N | 328 | 292 | 260 |
| Boosted | 87 (26.5) | 73 (25.0) | 67 (25.8) |
| Primed | 86 (26.2) | 77 (26.4) | 72 (27.7) |
| Unvaccinated | 155 (47.3) | 142 (48.6) | 121 (46.5) |
| # Of Covid-19 Symptoms, Mean (Sd) | 3.1 (3.6) | 2.7 (3.5) | 2.6(3.7) |
| **Symptom Description [SHORT NAME]** | **n (%)** | **n (%)** | **n (%)** |
| Tiredness or fatigue that interferes with daily life [TIRED] | 135 (41%) | 108 (37%) | 101 (39%) |
| Symptoms that get worse after physical or mental effort [WORSE] | 50 (15%) | 36 (12%) | 28 (11%) |
| Chest pain [PAIN] | 50 (15%) | 43 (15%) | 36(14%) |
| Difficulty breathing or shortness of breath SOB | 58(18%) | 42 (14%) | 32(12%) |
| Cough [COUGH] | 86(26%) | 38(13%) | 32(12%) |
| Chest pain [CHEST] | 32(10%) | 17(6%) | 17(7%) |
| Fast-beating or pounding heart [HEART] | 38(12%) | 29(10%) | 29(11%) |
| Change in smell or taste [TASTE] | 51(16%) | 35(12%) | 27(10%) |
| Headache [HEADACHE] | 67(20%) | 51(17%) | 50(19%) |
| Dizziness when you stand up [DIZZY] | 45(14%) | 43(15%) | 38(15%) |
| Difficulty thinking or concentrating [FOG] | 86(26%) | 70(24%) | 58(22%) |
| Pins-and-needles feelings [PINS] | 24(7%) | 28(10%) | 27(10%) |
| Sleep problems [SLEEP] | 81(25%) | 63(22%) | 52(20%) |
| Depression or anxiety [MOOD] | 36(11%) | 31(11%) | 32(12%) |
| Problems with memory [MEMORY] | 38(12%) | 35(12%) | 14(5%) |
| Diarrhea [DIARRHEA] | 23(7%) | 18(6%) | 14(5%) |
| Joint or muscle pain [MUSCLE] | 66(20%) | 53(18%) | 38(15%) |
| Rash [RASH] | 11(3%) | 9(3%) | 5(2%) |
| Activity Impairment Mean (Sd) | 20.5 (25.6) | 19.5 (26.0) | 18.6 (25.4) |
| HRQol Utility Index, Mean (Sd) | 0.86 (0.17) | 0.86 (0.20) | 0.86 (0.28) |

We explored and evaluated our LCA model according to best practices (Weller, Bowen, and Faubert 2020) by assessing the relationship between predicted Latent Class and the known patient outcomes 1) Activity Impairment, 2) Health-Related Quality of Life, and 3) Number of symptoms. Finally, we calculated the odds ratio of Latent Class by vaccination status.

## Results

A total of 676 symptomatic SARS-CoV-2 subjects were consented during the acute stage, but only 328 (49%) completed first PASC symptom survey at 4 weeks and were included in this study. Of these 328, about 26% of study participants were primed, 26% were boosted, and 47% were unvaccinated at Week 4. By the end of the study period, 68 (21%) participants were lost to follow-up, resulting in 260 participants completing the final survey at Month 6.

At week 4, participants reported an average of 3.1 PASC related symptoms, with symptoms declining gradually to 2.6 by Month 6. The most common symptoms at week 4 were tiredness (41%), cough (26%), difficulty thinking (26%), and difficulty sleeping (25%). Except for cough, which declined rapidly, the three most common symptoms remained somewhat stable over month 3 and month 6.

### Latent Classes of Post-COVID Symptoms

Models having between 2 to 10 latent classes were evaluated for use. We present 2 through 5 class models for reference. Generally, lower Bayesian Information Criterion (BIC) and Akaike Information Criterion (AIC) indicate better model fit.

Results from the LCA suggest that latent classes of PASC symptoms exist for all the timepoints (up to six months). As shown in Table 2, the BIC suggested a three-class model. Although entropy (a measure of class separation) was not used to select a final model, it is worth noting that the three-class model had adequate entropy>.80 (Larose et al. 2016).

**Table 2:** Number of Latent Classes & Model Fit Criteria.

|  |  | 2-Class | 3-Class | 4-Class | 5-Class |
| --- | --- | --- | --- | --- | --- |
| Week 4 | AIC | 4128.87 | 4058.92 | 4056.90 | 4051.76 |
|  | BIC | 4276.80 | 4282.71 | 4356.55 | 4427.27 |
| Month 3 | AIC | 3409.29 | 3289.39 | 3289.56 | 3292.08 |
|  | BIC | 3552.69 | 3506.32 | 3580.03 | 3656.08 |
| Month 6 | AIC | 2875.81 | 2770.79 | 2751.48 | 2757.33 |
|  | BIC | 3014.67 | 2980.87 | 3032.78 | 3109.84 |

**Error! Reference source not found.** shows a graphical representation of the three-class model. The x-axis lists the names of the PASC self-reported symptoms. The y-axis indicates the probability of class membership for each of the symptoms. Therefore, probabilities close to 1 suggest greater symptom representation and subsequent worst health condition (Figure 1).

**Figure 1:**
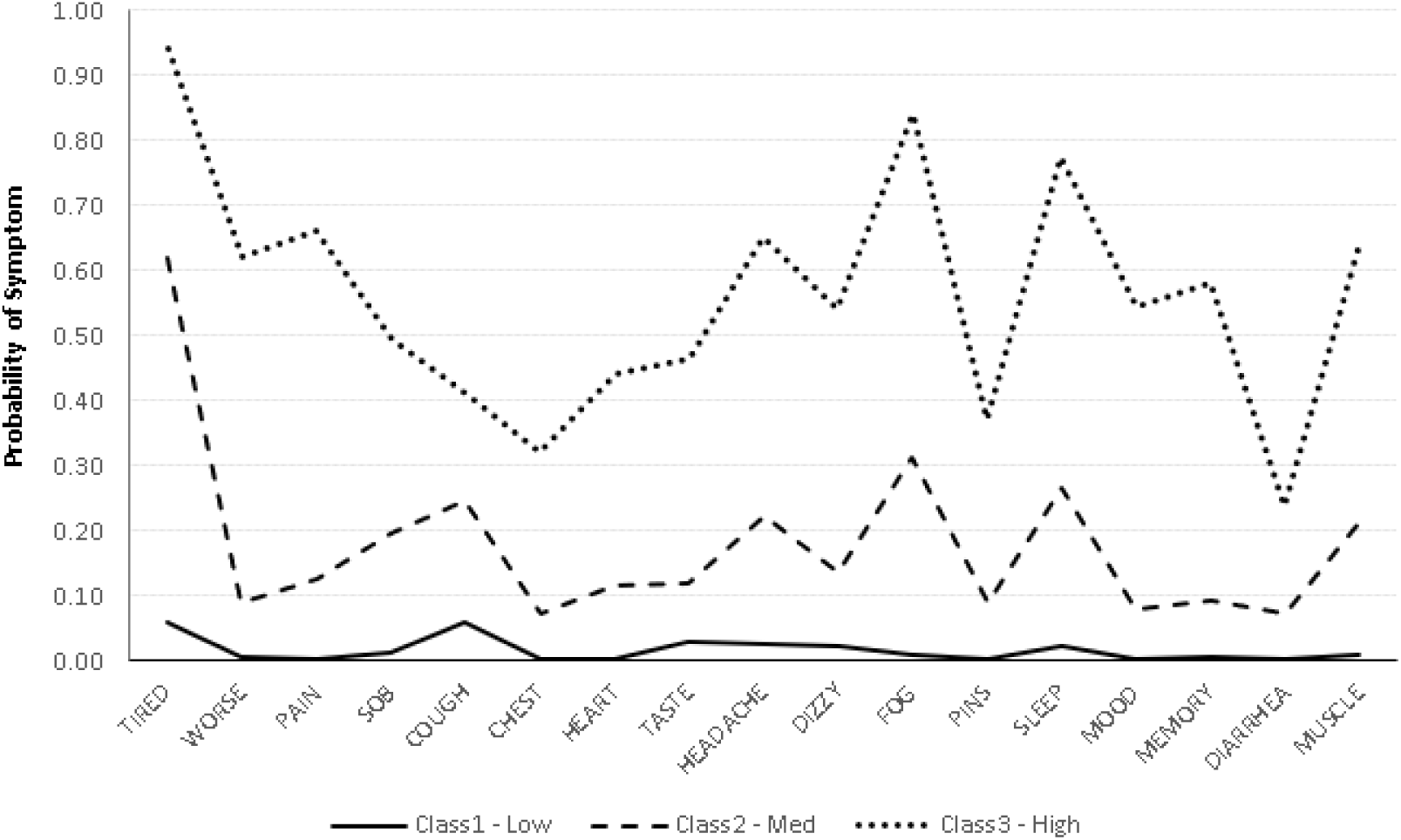
Latent Class Profiles of PASC Symptoms Suggest Symptom Burden.

When the selected 3-class model is plotted (Figure 1), the three classes form roughly parallel lines and do not appear to represent different ‘mixtures’ of symptoms. Similar LCA patterns have been suggested to represent severity or burden of the disease (Sinha, Calfee, and Delucchi 2021), rather than distinct subtypes. Accordingly, we interpret the three classes as levels of Symptom Burden: *Low*, *Medium,* and *High Symptom Burden*.

**Error! Reference source not found.** also illustrates the basic characteristics of the three classes based on the 17 symptoms. The majority (50%) of our symptomatic, SARS-CoV-2 positive adults were in the *Medium Symptom Burden* class. *High Symptom Burden* and *Low Symptom Burden* classes contained fifteen (15%) and thirty-five (35%) percent of our participants, respectively. For further characterization of our latent classes, see Appendix Table A-1 which shows the probability of symptoms reported given a latent class group, and Appendix Figure A-1 which shows the participant movement into and out of classes over our time periods.

### Latent Classes and Outcomes

Figure 2 plots the latent Symptom Burden classes over three time points: 4 Weeks, 3 Months, and 6 Months. Figure 3 shows the relationship between Latent Symptom Burden Class and reported activity impairment. Figure 4 shows the relationship between Latent Symptom Burden Class and reported Quality of Life. Figure 5 shows the relationship between Latent Symptom Burden Class and number of symptoms.

**Figure 2:**
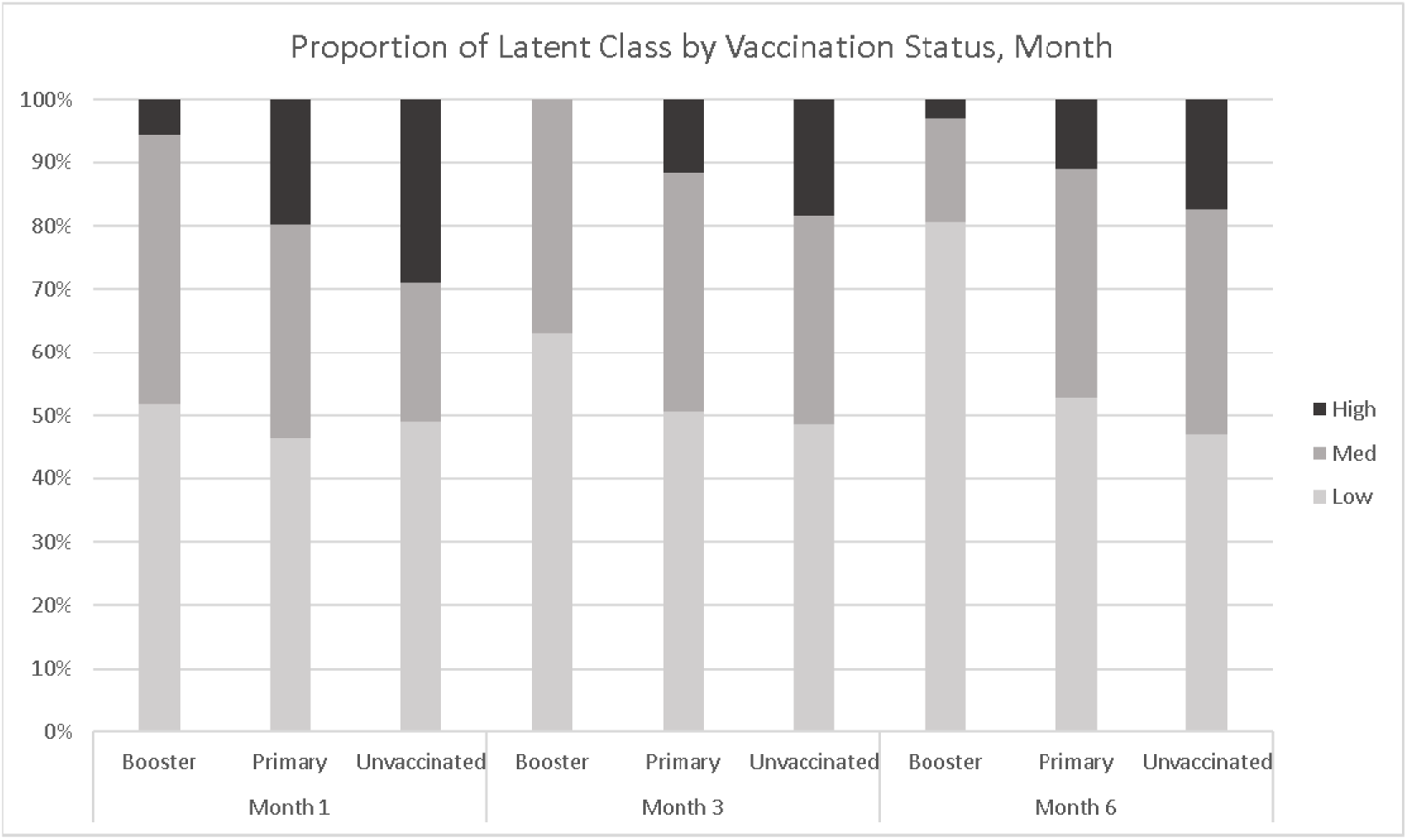
Latent Symptom Burden Class Distribution Over Three Time Points.

**Figure 3:**
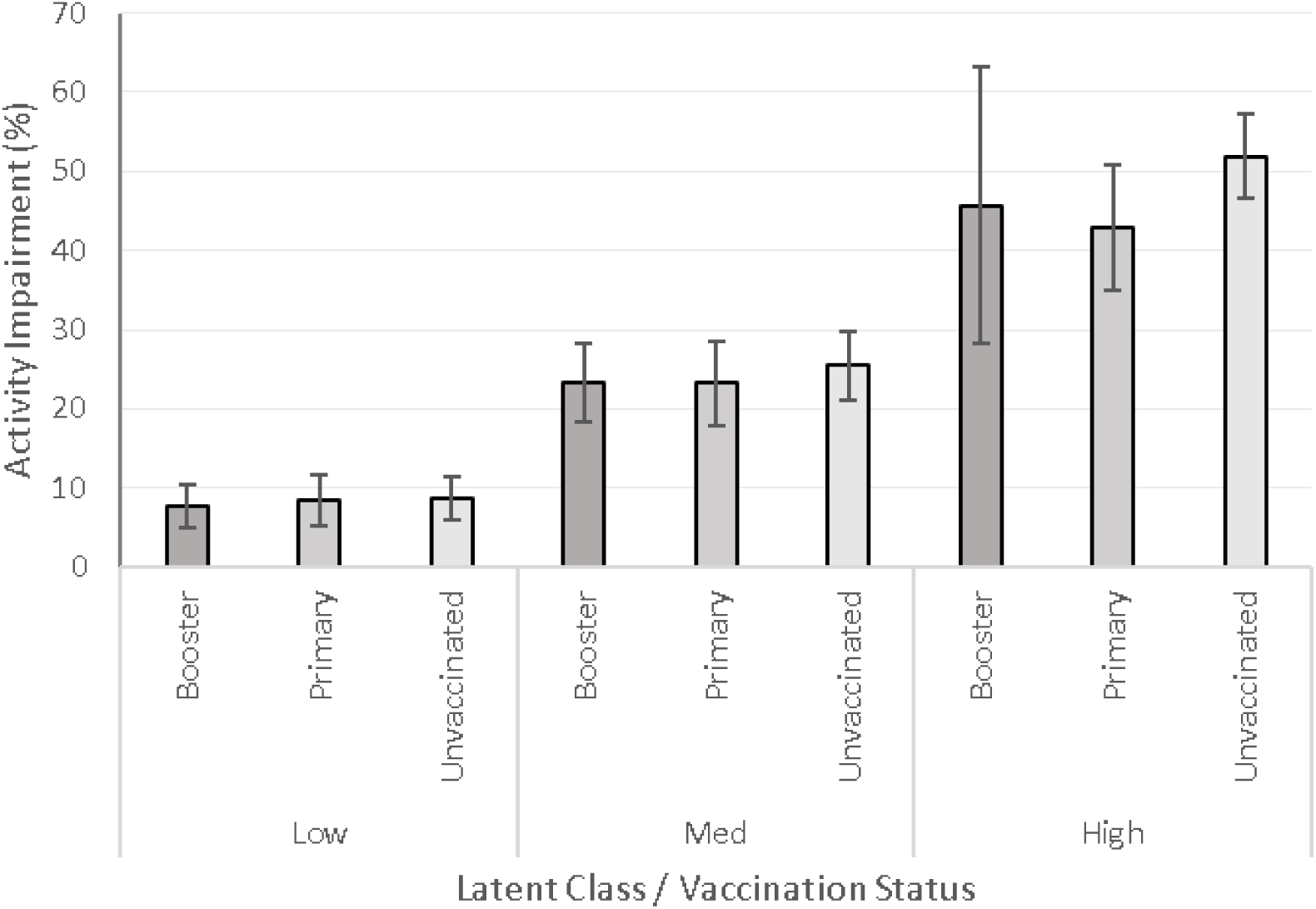
Percentage of Reported Activity Impairment by Latent Symptom Burden Class.

**Figure 4:**
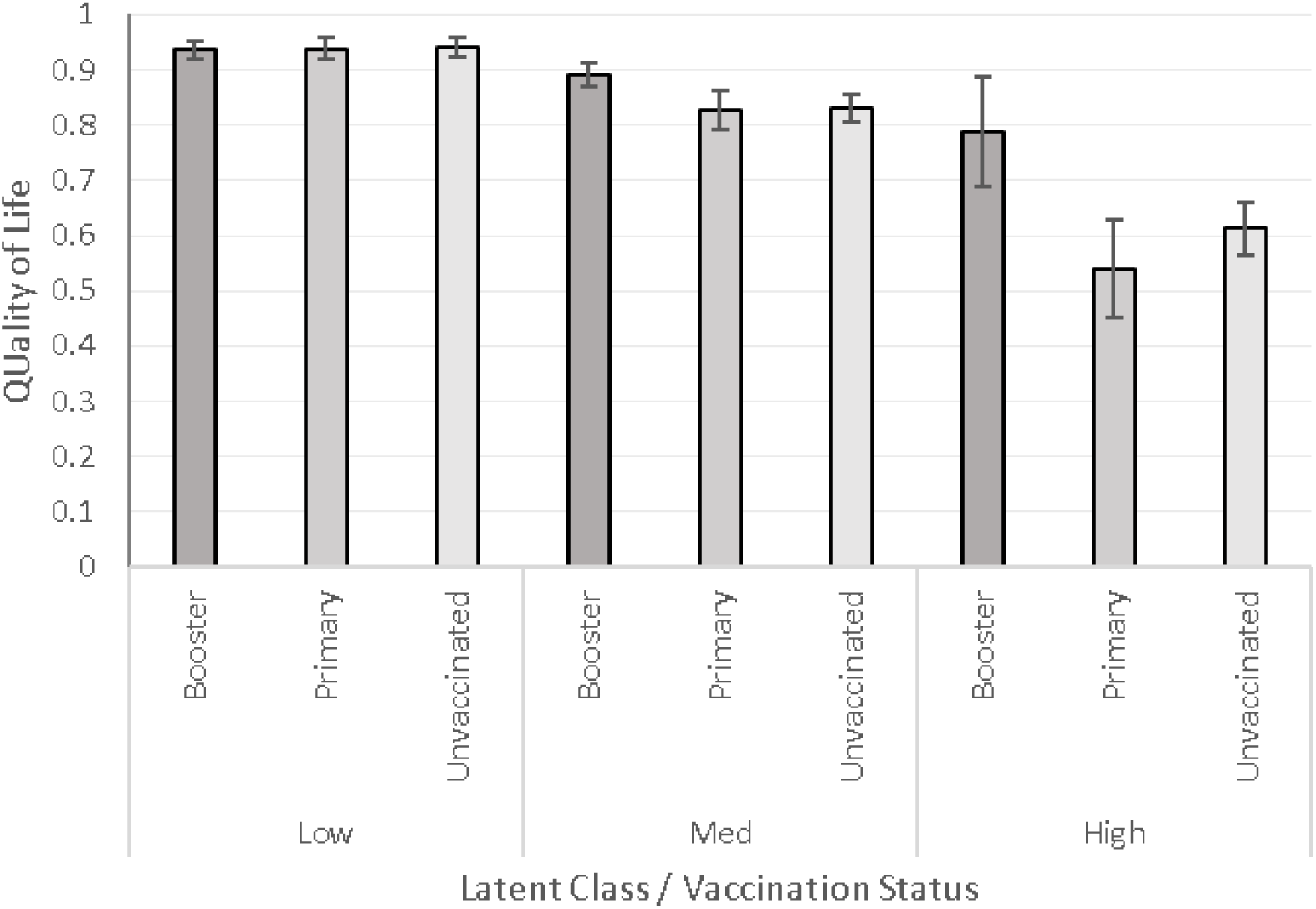
Quality of Life by Latent Symptom Burden Class.

**Figure 5:**
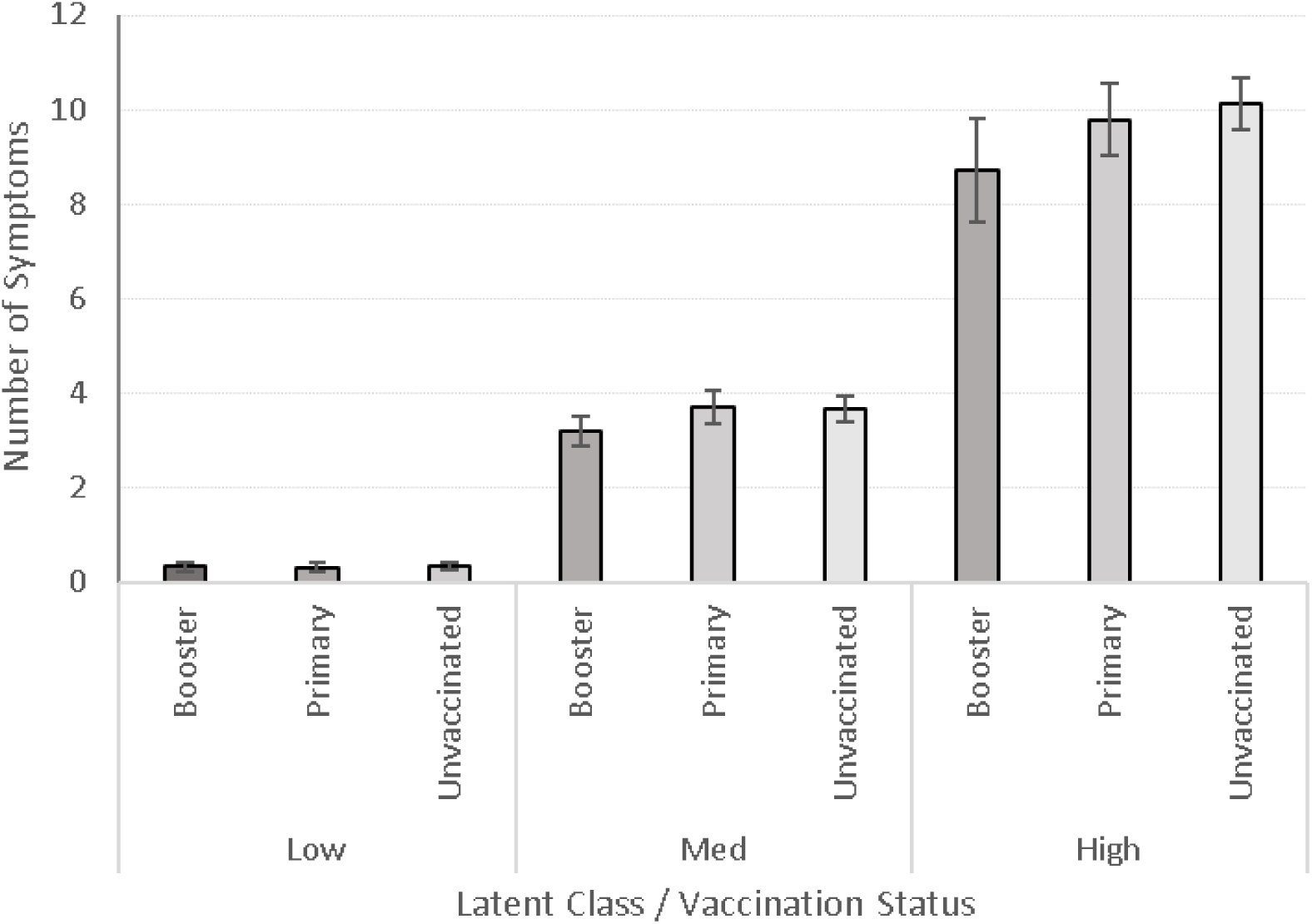
Number of Symptoms by Latent Symptom Burden Class.

#### Impact of Sars-Cov-2 Vaccination on Latent Symptom Burden Class

We also explored our LCA model by testing the association of vaccination status to latent class (Table 3). Compared to unvaccinated, individuals with the primary vaccination series were less likely to be in the High Symptom Burden class. Boosted vaccination appears to have a stronger association with latent class. Individuals with a boosted vaccination were about twice as likely to be in the Low Symptom Burden class, less likely to be in Medium Symptom Burden class, and much less likely to be in the High Symptom Burden class.

**Table 3.** Odds Ratio of Latent Symptom Burden Class by Vaccination Status.

|  | Low Burden |  | Medium Burden |  | High Burden |  |
| --- | --- | --- | --- | --- | --- | --- |
| Ref = Un- vaccinated | OR | 95% CI | OR | 95% CI | OR | 95% CI |
| Primary | 1.23 | 0.89-1.70 | 1.24 | 0.88-1.73 | 0.61 | 0.39- 0.92 |
| Boosted | 2.24 | 1.61- 3.15 | 0.77 | 0.79-0.77 | 0.08 | 0.03- 0.18 |

## Discussion

The result of this work suggests that LCA can be used to identify subgroups of PASC based on self-repored symtoms. Our LCA suggests three latent classes that we interpret as *Low Symptom Burden*, *Medium Symptom Burden*, and *High Symptom Burden* PASC. Vaccinated individuals were more likely to be in the *Low Symptom Burden* latent classes at all time points compared to unvaccinated individuals, up to 6 months. This finding is relevant to the question of how long SARS-CoV-2 vaccines may be protective (Tenforde, Link-Gelles, and Patel 2022), considering the subjects’ most recent vaccination may be months old prior to enrollment in our study. As previously reported, individuals in the primary vaccine group were vaccinated up to 11 months before the study (Di Fusco et al. 2023). In the more recently available booster group, some were boosted up to 4 months before the study (Di Fusco et al. 2023). Reducing the number of SARS-COV-2 symptoms is associated with reduced risk of PASC (Gao, Liu, and Liu 2022).

### Compared to Prior Work

To our knowledge, this is the first research to examine the latent classes of laboratory confirmed SARS-COV-2 symptoms over a six-month period in association with PASC. It is useful to consider these results within the context of similar studies. Yet, to date, only two studies have applied LCA to the spectrum of SARS-COV-2 symptoms. When conducting LCA on symptomatic, SARS-CoV-2 -unconfirmed patients, Hosseinzadeh and colleagues also identified three latent classes of symptoms during acute SARS-CoV-2 infection (Hosseinzadeh et al. 2021). Unlike our study, Hosseinzadeh included SARS-CoV-2 unconfirmed patients and applied the LCA classes to predict a positive SARS-CoV-2 PCR test.

Moreira and Colleagues also conducted LCA on symptoms during acute SARS-CoV-2 infection without laboratory confirmed SARS-CoV-2 status in participants. Moreira’s LCA model resulted in six (6) latent classes: (1) all symptoms; (2) high prevalence of symptoms; (3) predominance of fever; (4) predominance of cough/sore throat; (5) mild symptoms with predominance of headache; and (6) absence of symptoms (Moreira 2021).

Although these two LCA studies explored symptoms during acute SARS-CoV-2 infection, their results are comparable to our analysis. All three LCA class models resulted in general levels of burden of symptoms. However unlike our study where no class favored one symptom over another, Moreira and Hosseinzadeh found that symptoms of fever (both), cough/sore throat (both), headache (Moreira only), and sore throat (Hosseinzadeh only) behaved independently of the rest (Hosseinzadeh et al. 2021, Moreira 2021). It is possible that these differences can be attributed to individuals with another clinical course that was not SARS-CoV-2. This notion is supported by Hosseinzadeh’s finding that “sore throat” dropped from the “*most likely to be COVID PCR+*” class while all other symptoms remained.

This analysis supports earlier work that found that the BNT162b2 booster could reduce the risk and burden of PASC (Di Fusco et al. 2023). Boosted participants reported fewer and less durable symptoms and vaccination was associated with lower latent symptom burden class, increased quality of life, and decreased activity impairment (Di Fusco et al. 2023).

### Limitations

LCA models, particularly those that result in nested classes like ours, must be interpreted prudently. It is possible that our LCA regressed to a simpler k-means model (Sinha, Calfee, and Delucchi 2021). If this were the case, the model would represent equal segments of the distribution of symptoms. However, our fit statistics indicate that 3 classes are a better fit than 2 or 4, suggesting that 3 classes are distinct, and 2 or 4 classes are less distinct. Additionally, exploratory anlaysis supported our model. We may also be misinterpreting the classes. Although our evaluations support our interpretation, it is possible that class 1, class 2, and class 3, represent hidden variables entirely different than *Low*, *Medium*, and *High Symptom Burden*. Another limitation is that we defined the clusters based on the presence of symptoms, which is only one way to look at impact and symptomatology of a condition. Further research is warranted to look into severity of symptoms rather than presence. Athough we selected our validation comparators (# of symptoms, activity impairment, and Quality of Life) a priori, we do not know how the model would fare with other comparators.

## Conclusion

Latent Class Analysis is growing in popularity as a robust method to cluster symptoms for acute respiratory conditions such as SARS-CoV-2 infection. Our LCA identified three PASC phenotypes: *Low Symptom Burden*, *Medium Symptom Burden*, and *High Symptom Burden*. This model had high fit statistics, remained stable over our three time periods, and was statistically associated with vaccination status.

Further validation analysis of the model revealed that *Low Symptom Burden* was associated with lower activity impairment, higher quality of life, and a lower number of PASC related symptoms, compared to the *High Symptom Burden* class.

Although not the focus of this methodological paper, our analysis supports the prior work of (Hosseinzadeh et al. 2021, Moreira 2021) and suggests LCA may perform as adequate predictors of diagnosis and prognosis for PASC related symptoms.

## DECLARATIONS

## Ethics Approval and consent to participate

This study was approved by the Sterling IRB, Protocol #C4591034. Participation in the study was voluntary and anonymous. Consent was obtained electronically via the CVS Health E-Consent platform. Participants were informed of their right to refuse or withdraw from the study at any time. Participants were compensated for their time.

## Consent for publication

All authors have given their approval for this manuscript version to be published.

## Availability of data and material

Aggregated data that support the findings of this study are available upon reasonable request from the corresponding author XS, subject to review. These data are not publicly available due to them containing information that could compromise research participant privacy/consent.

## Authors’ contributions

All named authors meet the International Committee of Medical Journal Editors (ICMJE) criteria for authorship for this article. All authors contributed to study conception and design, data acquisition, analysis, and interpretation, drafting and revising of the manuscript.

## Acknowledgements

We would like to acknowledge Shiyu Lin and Leena Samuel of CVS Health for their analytic contributions and editorial support, respectively.

## APPENDIX

**TABLE A1:** Probability of Symptoms Given Latent Class (class interpretation)

|  | **CLASS 3** | **CLASS 2** | **CLASS 1** |
| --- | --- | --- | --- |
|  | **(HIGH)** | **(MEDIUM)** | **(LOW)** |
| **TIRED** | 0.94 | 0.62 | 0.06 |
| **WORSE** | 0.62 | 0.09 | 0.00 |
| **PAIN** | 0.66 | 0.13 | 0.00 |
| **SOB** | 0.49 | 0.19 | 0.01 |
| **COUGH** | 0.41 | 0.24 | 0.06 |
| **CHEST** | 0.32 | 0.07 | 0.00 |
| **HEART** | 0.44 | 0.12 | 0.00 |
| **TASTE** | 0.46 | 0.12 | 0.03 |
| **HEADACHE** | 0.65 | 0.22 | 0.02 |
| **DIZZY** | 0.54 | 0.14 | 0.02 |
| **FOG** | 0.84 | 0.31 | 0.01 |
| **PINS** | 0.37 | 0.09 | 0.00 |
| **SLEEP** | 0.77 | 0.26 | 0.02 |
| **MOOD** | 0.55 | 0.08 | 0.00 |
| **MEMORY** | 0.58 | 0.09 | 0.01 |
| **DIARRHEA** | 0.24 | 0.07 | 0.00 |
| **MUSCLE** | 0.64 | 0.21 | 0.01 |

**Figure A1:**
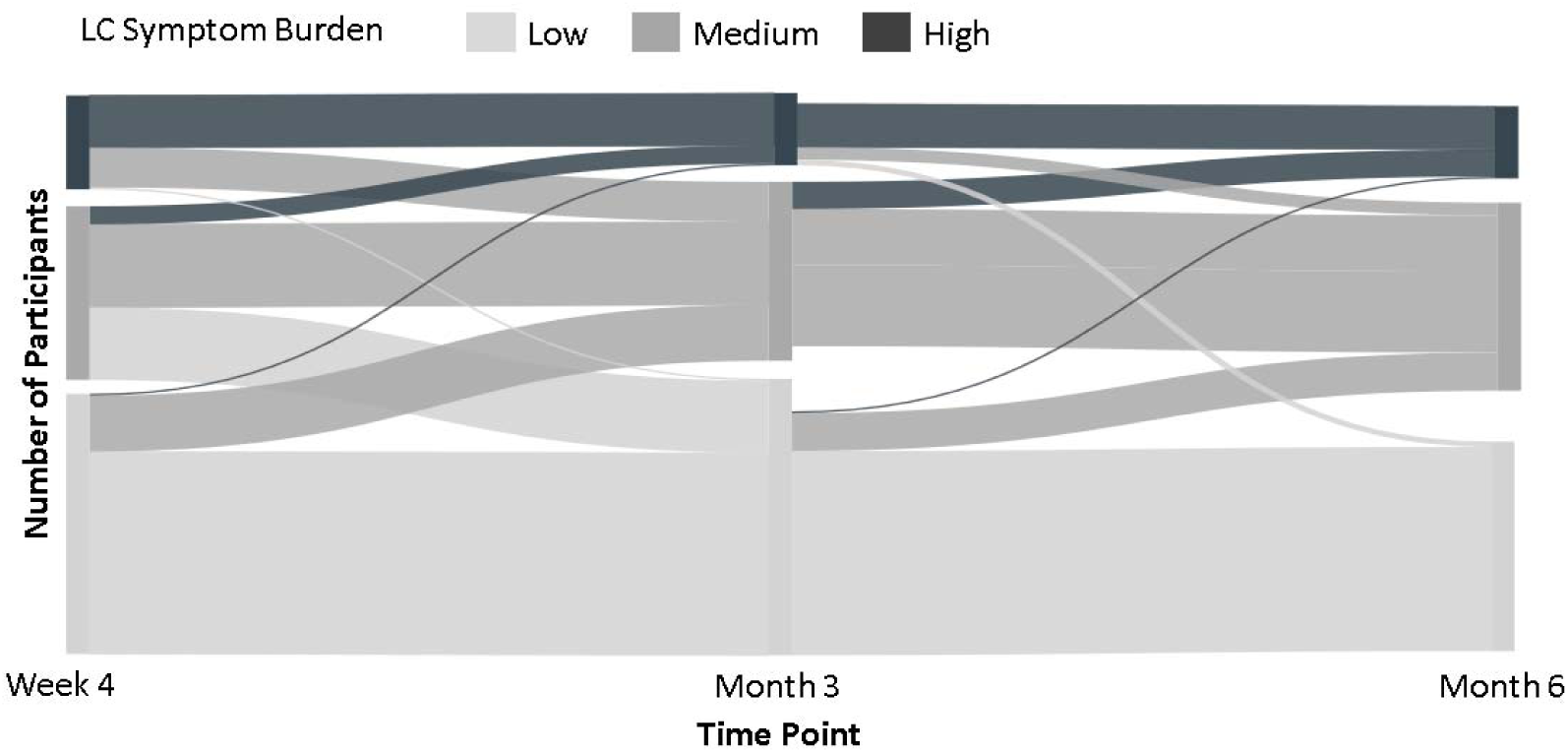
Class Membership (Symptom Burden) Changes Over Time. This Sankey chart shows how class membership (Symptom Burden) changes over the three time points. Most move to lower burden over time. A small number move up in increased burden over time.

## References

Asparouhov T, Muthén B 2014. Auxiliary variables in mixture modeling: Three-step approaches using Mplus. 21: 329–341. doi:10.1080/10705511.2014.915181.

Atreya, M. R., L. N. Sanchez-Pinto, and R. Kamaleswaran. 2022. “Commentary: ‘Critical illness subclasses: all roads lead to Rome’.” Crit Care 26 (1):387. doi: 10.1186/s13054-022-04265-w.

Calfee, C. S., K. Delucchi, P. E. Parsons, B. T. Thompson, L. B. Ware, and M. A. Matthay. 2014. “Subphenotypes in acute respiratory distress syndrome: latent class analysis of data from two randomised controlled trials.” Lancet Respir Med 2 (8):611–20. doi: 10.1016/s2213-2600(14)70097-9.

Centers for Disease Control and Prevention. 2022. Long COVID or Post-COVID Conditions.

Dahmer, Mary K, Guangyu Yang, Min Zhang, Michael W Quasney, Anil Sapru, Heidi M Weeks, Pratik Sinha, Martha AQ Curley, Kevin L Delucchi, and Carolyn S Calfee. 2022. “Identification of phenotypes in paediatric patients with acute respiratory distress syndrome: a latent class analysis.” The Lancet Respiratory Medicine 10 (3):289–297.

Di Fusco, M., X. Sun, M. M. Moran, H. Coetzer, J. M. Zamparo, L. Puzniak, M. B. Alvarez, Y. P. Tabak, and J. C. Cappelleri. 2022. “Impact of COVID-19 and effects of BNT162b2 on patient-reported outcomes: quality of life, symptoms, and work productivity among US adult outpatients.” J Patient Rep Outcomes 6 (1):123. doi: 10.1186/s41687-022-00528-w.

Di Fusco, Manuela, Xiaowu Sun, Mary M. Moran, Henriette Coetzer, Joann M. Zamparo, Mary B. Alvarez, Laura Puzniak, Ying P. Tabak, and Joseph C. Cappelleri. 2023. “Impact of COVID-19 and effects of booster vaccination with BNT162b2 on six-month long COVID symptoms, quality of life, work productivity and activity impairment during Omicron.” medRxiv:2023.03.15.23286981. doi: 10.1101/2023.03.15.23286981.

EuroQol Research Foundation. 2019. EQ-5D-5L User Guide, Version 3.0.

Famous, K. R., K. Delucchi, L. B. Ware, K. N. Kangelaris, K. D. Liu, B. T. Thompson, and C. S. Calfee. 2017. “Acute Respiratory Distress Syndrome Subphenotypes Respond Differently to Randomized Fluid Management Strategy.” Am J Respir Crit Care Med 195 (3):331–338. doi: 10.1164/rccm.201603-0645OC.

French, N., G. Jones, C. Heuer, V. Hope, S. Jefferies, P. Muellner, A. McNeill, S. Haslett, and P. Priest. 2021. “Creating symptom-based criteria for diagnostic testing: a case study based on a multivariate analysis of data collected during the first wave of the COVID-19 pandemic in New Zealand.” BMC Infect Dis 21 (1):1119. doi: 10.1186/s12879-021-06810-4.

Gao, P., J. Liu, and M. Liu. 2022. “Effect of COVID-19 Vaccines on Reducing the Risk of Long COVID in the Real World: A Systematic Review and Meta-Analysis.” Int J Environ Res Public Health 19 (19). doi: 10.3390/ijerph191912422.

Hosseinzadeh, A., M. Rezapour, M. Rohani-Rasaf, M. H. Emamian, S. Talebi, S. Goli, R. Chaman, H. Sheibani, E. Binesh, F. Zare, and A. Khosravi. 2021. “Epidemiological patterns of syndromic symptoms in suspected patients with COVID-19 in Iran: A Latent Class Analysis.” J Res Health Sci 21 (1):e00508. doi: 10.34172/jrhs.2021.41.

Karadaş, Ö, B. Öztürk, A. R. Sonkaya, B. Taşdelen, A. Özge, and H. Bolay. 2021. “Latent class cluster analysis identified hidden headache phenotypes in COVID-19: impact of pulmonary infiltration and IL-6.” Neurol Sci 42 (5):1665–1673. doi: 10.1007/s10072-020-04978-2.

Larose, C., O. Harel, K. Kordas, and D. K. Dey. 2016. “Latent Class Analysis of Incomplete Data via an Entropy-Based Criterion.” Stat Methodol 32:107–121. doi: 10.1016/j.stamet.2016.04.004.

Masyn, Katherine E. 2013. “Latent class analysis and finite mixture modeling.” In The Oxford handbook of quantitative methods: Statistical analysis, Vol. 2, 551–611. New York, NY, US: Oxford University Press.

Moreira, R. D. S. 2021. “Latent class analysis of COVID-19 symptoms in Brazil: results of the PNAD-COVID19 survey.” Cad Saude Publica 37 (1):e00238420. doi: 10.1590/0102-311x00238420.

Nalbandian, A., K. Sehgal, A. Gupta, M. V. Madhavan, C. McGroder, J. S. Stevens, J. R. Cook, A. S. Nordvig, D. Shalev, T. S. Sehrawat, N. Ahluwalia, B. Bikdeli, D. Dietz, C. Der-Nigoghossian, N. Liyanage-Don, G. F. Rosner, E. J. Bernstein, S. Mohan, A. A. Beckley, D. S. Seres, T. K. Choueiri, N. Uriel, J. C. Ausiello, D. Accili, D. E. Freedberg, M. Baldwin, A. Schwartz, D. Brodie, C. K. Garcia, M. S. V. Elkind, J. M. Connors, J. P. Bilezikian, D. W. Landry, and E. Y. Wan. 2021. “Post-acute COVID-19 syndrome.” Nat Med 27 (4):601–615. doi: 10.1038/s41591-021-01283-z.

Natarajan, A., A. Shetty, G. Delanerolle, Y. Zeng, Y. Zhang, V. Raymont, S. Rathod, S. Halabi, K. Elliot, J. Q. Shi, and P. Phiri. 2023. “A systematic review and meta-analysis of long COVID symptoms.” Syst Rev 12 (1):88. doi: 10.1186/s13643-023-02250-0.

Nylund, Karen L., Tihomir Asparouhov, and Bengt O. Muthén. 2007. “Deciding on the Number of Classes in Latent Class Analysis and Growth Mixture Modeling: A Monte Carlo Simulation Study.” Structural Equation Modeling: A Multidisciplinary Journal 14 (4):535–569. doi: 10.1080/10705510701575396.

Pickard, A. Simon, Ernest H. Law, Ruixuan Jiang, Eleanor Pullenayegum, James W. Shaw, Feng Xie, Mark Oppe, Kristina S. Boye, Richard H. Chapman, Cynthia L. Gong, Alan Balch, and Jan J. V. Busschbach. 2019. “United States Valuation of EQ-5D-5L Health States Using an International Protocol.” Value in Health 22 (8):931–941. doi: 10.1016/j.jval.2019.02.009.

Priyal, V. Sehgal, S. Kapila, R. Taneja, P. Mehmi, and N. Gulati. 2023. “Review of Neurological Manifestations of SARS-CoV-2.” Cureus 15 (4):e38194. doi: 10.7759/cureus.38194.

Reilly, M. C., A. S. Zbrozek, and E. M. Dukes. 1993. “The validity and reproducibility of a work productivity and activity impairment instrument.” Pharmacoeconomics 4 (5):353–65. doi: 10.2165/00019053-199304050-00006.

Sigwadhi, L. N., J. L. Tamuzi, A. E. Zemlin, Z. C. Chapanduka, B. W. Allwood, C. F. Koegelenberg, E. M. Irusen, U. Lalla, V. D. Ngah, A. Yalew, P. Savieri, I. Fwemba, T. P. Jalavu, R. T. Erasmus, T. E. Matsha, A. Zumla, and P. S. Nyasulu. 2022. “Latent class analysis: an innovative approach for identification of clinical and laboratory markers of disease severity among COVID-19 patients admitted to the intensive care unit.” IJID Reg 5:154–162. doi: 10.1016/j.ijregi.2022.10.004.

Sinha, P., C. S. Calfee, and K. L. Delucchi. 2021. “Practitioner’s Guide to Latent Class Analysis: Methodological Considerations and Common Pitfalls.” Crit Care Med 49 (1):e63–e79. doi: 10.1097/ccm.0000000000004710.

Sinha, P., D. Furfaro, M. J. Cummings, D. Abrams, K. Delucchi, M. V. Maddali, J. He, A. Thompson, M. Murn, J. Fountain, A. Rosen, S. Y. Robbins-Juarez, M. A. Adan, T. Satish, M. Madhavan, A. Gupta, A. K. Lyashchenko, C. Agerstrand, N. H. Yip, K. M. Burkart, J. R. Beitler, M. R. Baldwin, C. S. Calfee, D. Brodie, and M. R. O’Donnell. 2021. “Latent Class Analysis Reveals COVID-19-related Acute Respiratory Distress Syndrome Subgroups with Differential Responses to Corticosteroids.” Am J Respir Crit Care Med 204 (11):1274–1285. doi: 10.1164/rccm.202105-1302OC.

Tenforde, Mark W., Ruth Link-Gelles, and Manish M. Patel. 2022. “Long-term Protection Associated With COVID-19 Vaccination and Prior Infection.” JAMA 328 (14):1402–1404. doi: 10.1001/jama.2022.14660.

Thaweethai, Tanayott, Sarah E. Jolley, Elizabeth W. Karlson, Emily B. Levitan, Bruce Levy, Grace A. McComsey, Lisa McCorkell, Girish N. Nadkarni, Sairam Parthasarathy, Upinder Singh, Tiffany A. Walker, Caitlin A. Selvaggi, Daniel J. Shinnick, Carolin C. M. Schulte, Rachel Atchley-Challenner, Leora I. Horwitz, Andrea S. Foulkes, RECOVER Consortium Authors, and RECOVER Consortium. 2023. “Development of a Definition of Postacute Sequelae of SARS-CoV-2 Infection.” JAMA. doi: 10.1001/jama.2023.8823.

van Kessel, S. A. M., T. C. Olde Hartman, Plbj Lucassen, and C. H. M. van Jaarsveld. 2022. “Post-acute and long-COVID-19 symptoms in patients with mild diseases: a systematic review.” Fam Pract 39 (1):159–167. doi: 10.1093/fampra/cmab076.

Vermunt, Jeroen K. 2010. “Latent Class Modeling with Covariates: Two Improved Three-Step Approaches.” Political Analysis 18 (4):450–469.

Wang, X., L. Jehi, X. Ji, and P. J. Mazzone. 2021. “Phenotypes and Subphenotypes of Patients With COVID-19: A Latent Class Modeling Analysis.” Chest 159 (6):2191–2204. doi: 10.1016/j.chest.2021.01.057.

Weller, Bridget E., Natasha K. Bowen, and Sarah J. Faubert. 2020. “Latent Class Analysis: A Guide to Best Practice.” Journal of Black Psychology 46 (4):287–311. doi: 10.1177/0095798420930932.

Yugar-Toledo, J. C., L. B. T. Yugar, L. G. Sedenho-Prado, R. Schreiber, and H. Moreno. 2023. “Pathophysiological effects of SARS-CoV-2 infection on the cardiovascular system and its clinical manifestations-a mini review.” Front Cardiovasc Med 10:1162837. doi: 10.3389/fcvm.2023.1162837.

